# Single-cell profiling identifies a CD8^bright^ CD244^bright^ Natural Killer cell subset that reflects disease activity in HLA-A29-positive *birdshot chorioretinopathy*

**DOI:** 10.1101/2022.09.11.22279821

**Authors:** Pulak R. Nath, Mary Maclean, Vijay Nagarajan, Jung Wha Lee, Mehmet Yakin, Aman Kumar, Hadi Nadali, Brian Schmidt, Koray D. Kaya, Shilpa Kodati, Alice Young, Rachel R. Caspi, Jonas J. W. Kuiper, H. Nida Sen

## Abstract

Birdshot chorioretinopathy uveitis (BCR-UV) is strongly associated with HLA-A29 which implicates MHC-I pathway mediated perturbation of natural killer (NK) cells as a potential disease mechanism. We profiled blood NK cells at single-cell resolution in a cohort of patients and healthy controls and investigated the links between NK cell subpopulations and disease activity. Flow cytometry analysis of major immune cell lineages revealed substantial expansion of the CD56^dim^ CD16+ NK cells in BCR-UV compared to healthy controls and to other types of non-infectious uveitis. *Ex vivo* restimulation showed that NK cells from BCR-UV patients exhibit increased secretion of TNF-alpha, a cytokine considered central to the pathogenesis of BCR-UV. Unbiased transcriptomic characterization at single-cell resolution established that the expanded CD16+ (i.e., *FCGR3A+*) NK cells also co-express high levels of *CD8A* and *CD244*, indicating expansion of a subset of CD56^dim^ CD16+ CD8+ NK cells in patients. Confirmation of these results by high-dimensional flow cytometry further showed that the BCR-UV-associated CD8^bright^ CD244^bright^ NK cells displayed activation receptors including CD314 (NKG2D), and cytotoxicity receptor CD337 (NKp30). Finally, longitudinal monitoring of patients showed that clinical remission after systemic immunomodulatory treatment correlated with a significant decrease in CD8^bright^ CD244^bright^ NK cells. In conclusion, there is an expansion of CD8^bright^ CD244^bright^ NK cells during active disease in BCR-UV patients which decrease upon successful systemic immunomodulatory treatment, suggesting that CD8^bright^/CD244^bright^ NK cells may be a pro-inflammatory NK subset involved in the underlying disease mechanism.

## INTRODUCTION

Non-infectious uveitis (NIU) is a clinically and prognostically heterogeneous group of ocular inflammatory diseases and a major cause of severe visual handicap^1^. Birdshot chorioretinopathy (Birdshot Uveitis or BCR-UV) is a relatively rare form of NIU that has clinically distinct features in the form of retinal and choroidal inflammatory lesions visible on examination. BCR-UV can lead to progressive deterioration of visual function^2, 3, 4^ due to persistent inflammation in the retina and choroid^5^. Consequently, patients often require systemic immunomodulatory therapy to control ocular inflammation^3, 6^. The disease mechanisms driving BCR-UV remain to be elucidated, but scientific advances have shed light on the immunopathology of this clinically well-defined ophthalmological condition^7^.

One of the most striking molecular features of BCR-UV is that it’s strongly associated with the presence of HLA-A29 allele such that all patients carry the HLA-A29 allele (most often the common *HLA-A*29:02*)^7, 8^. Genome-wide genetic studies have revealed that the susceptibility to BCR-UV also maps to other factors of the MHC-I pathway^9, 10, 11^. This incriminates CD8+ T cells and Natural Killer (NK) cells in the pathogenesis of BCR-UV^7^. Indeed, CD8+ T cells have been shown to infiltrate eye tissues of patients and secrete cytokines considered central to its immunopathology^12, 13, 14, 15^.

The role of NK cells in BCR-UV has remained significantly underexplored. Classically, blood NK cells are subdivided into two well-established overarching populations; the CD56^dim^ CD16+ NK cells (∼90% of circulating NK cells) also known as *NK1* cells which are considered to be cytotoxic and produce greater amounts of pro-inflammatory cytokines and the CD56^bright^ (∼10% of circulating NK cells) - or *NK2* cells - that are considered to be more immunoregulatory^16^. Of interest, CD56^bright^ NK cells have been shown to be lower in patients with NIU, including BCR-UV patients with active uveitis^17^ and successful immunosuppressive treatment of NIU is accompanied by the recovery of CD56^bright^ NK cell levels^18^. However, beyond this phenotypic bifurcation, NK cell population is substantially more diverse and thus far single-cell RNA sequencing (scRNAseq) has uncovered at least 10 transcriptionally distinct clusters in peripheral blood^19, 20, 21^. This includes ‘inflammatory’ CD56^dim^ CD16+ NK subsets that was defined by high levels of cytokine and interferon response genes^20, 21^ and “adaptive-like” NK populations that expand during infection^22^.

Transcriptomically distinct NK cell subsets are considered to exhibit differential effector functions mediated by an ensemble of surface immunoregulatory molecules^23^, in particular Killer cell immunoglobulin-like receptors (KIR), IgG Fc receptors (e.g., CD16), and integrins (e.g., CD47)^24, 25, 26^. Perturbations in the composition of the NK cells have been reported in other MHC-I associated conditions, such as HLA-B27-positive *ankylosing spondylitis*^27^ and were shown to be predictive for clinical outcome in autoimmune diseases, such as Multiple Sclerosis^28^. Collectively, these observations suggest that deep phenotyping of the blood NK cell compartment could provide better understanding of disease biology and may hold clinically relevant information to the clinical course of BCR-UV.

Here, we have taken a multi-omics approach to phenotype the NK cell compartment at single-cell resolution of patients with BCR-UV and report on the expansion of a CD56^dim^ CD16+ subset of NK cells which are CD8^bright^ and CD244^bright^, and whose reduction is correlated with clinical improvement after systemic immunosuppressive therapy.

## RESULTS

### Increased frequency of CD16+ NK cells in Birdshot uveitis patients

To investigate the relationship between NK cell-mediated inflammation and BCR-UV, we first sought evidence that circulating NK cells were specifically perturbed in BCR-UV. To this end, we quantified the major lineages of immune cells (10-marker panel, 6 lineages) (Supplementary Fig. 1A) in peripheral blood using flow cytometry in a cohort of 18 BCR-UV patients, 80 healthy controls, and 121 non-infectious uveitis (NIU) patients other than BCR-UV (Fig. 1A). Global comparison of major lineages in all NIU patients versus healthy controls revealed a significant increase in frequency of blood NK cells (Fig. 1B), but not T cells, B cells, monocytes, or dendritic cells (Supplementary Fig. 1B-D). Flow cytometry analysis revealed that blood NK cell frequency appeared to be increased in several uveitis subtypes, but this increase was most significant for BCR-UV (*P* < 0.0001) (Fig. 1C). This became more evident after quantification of the two major NK populations (i.e., NK1, and NK2) that can be distinguished by their expression of surface CD56 and CD16 (Fig. 1D). We detected a significant increase of CD56^dim^ CD16+ [NK1] cells and a concomitant decrease of the CD56^bright^ CD16^−^ [NK2] cells only in BCR-UV patients or when considering all NIU patients collectively, but not individually in any of the other types of NIU (Supplementary Fig. 1E-F). This skew in NK1/NK2 balance also remained evident after strict comparison to 15 age-matched healthy controls (mean age±SD = 62.2±8.8) (Fig. 1E).

**Figure 1.**
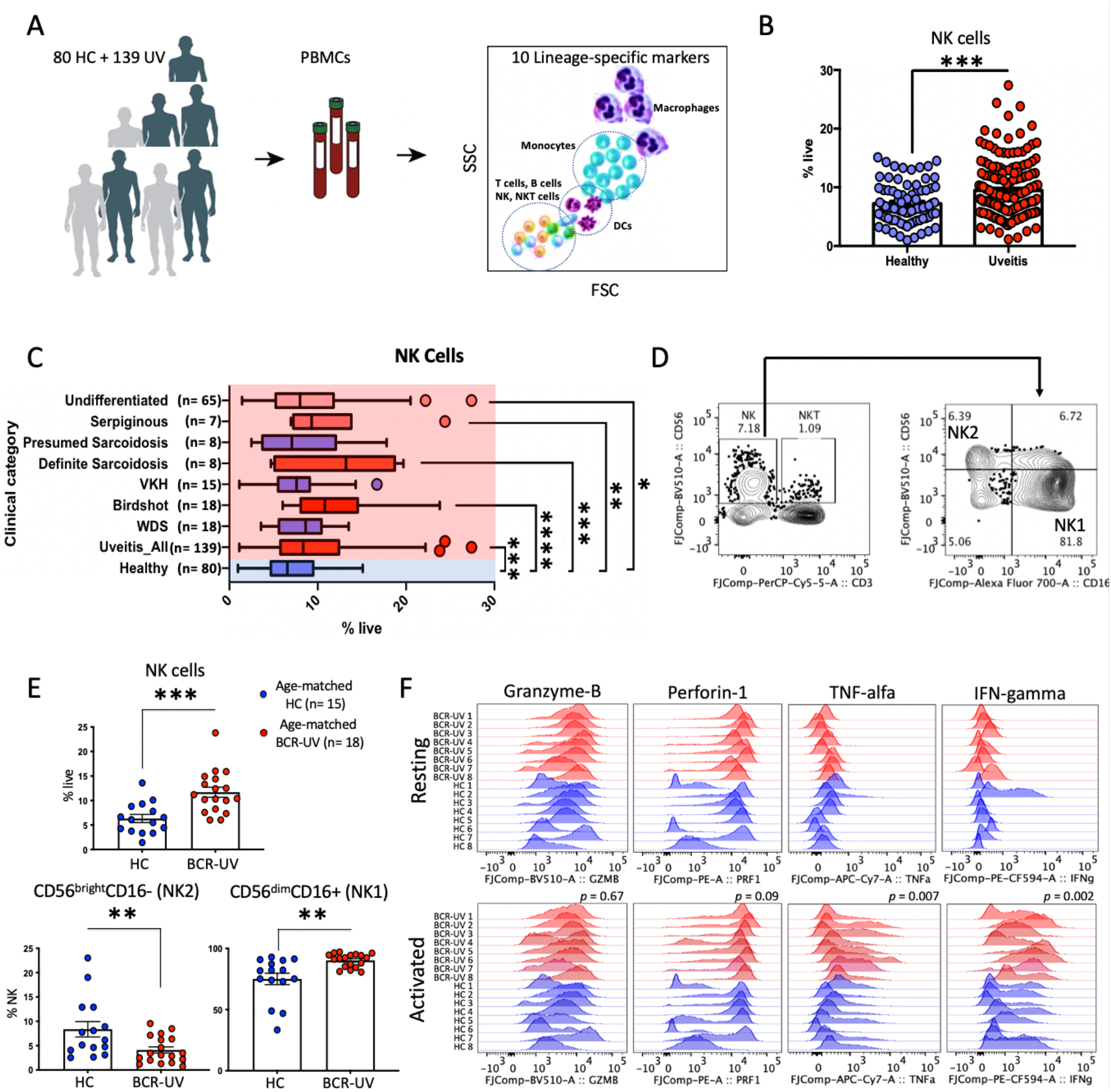
Flow-cytometry profiling revealed altered Natural killer (NK) cell composition in peripheral blood of Birdhshot Uveitis. **A.** Schematic representation of flow cytometry of major immune cell lineages in fresh peripheral blood of 139 uveitis patients (UV) and 80 age-, sex- matched healthy controls (HC). **B.** Flow-cytometry quantification of the percentage of NK cells in the peripheral blood of uveitis patients (Uveitis) compared to healthy controls (Healthy). **C.** Flow-cytometry analysis of NK cells (CD3^−^CD19^−^CD56^+^) in the fresh blood of different uveitis subgroups indicate the significant expansion of NK cells were restricted to birdshot, definite sarcoidosis, serpiginous and undifferentiated sub-groups of uveitis cohort. *P* values are from unpaired t test. **** *P* < 0.0001, *** *P* = 0.0002, ** *P* = 0.004, * *P* = 0.01. **D.** The flow-cytometry gating strategy for NK1 and NK2 subsets of NK cells using CD56 and CD16 in peripheral blood. **E.** NK cell and NK1 and NK2 subset quantification in peripheral blood of birdshot uveitis (BCR-UV) and age-matched healthy controls (HC). HC *n* = 15; BCR-UV *n* = 18. **F.** Histogram of the fluorescence intensity of intracellular effector molecules produced by NK cells and determined by flow cytometry analysis upon stimulation with Lymphocyte Activation Cocktail (BD Biosciences. Analysis was conducted using PBMCs from BCR-UV (red) and NK cells from healthy controls (HC, blue). HC *n* = 8; BCR-UV *n* = 8.

Importantly, NK cells of BCR-UV patients showed enhanced responsiveness to restimulation by production of significantly higher tumor necrosis factor-ɑ (TNF-ɑ, *P* = 0.007) and interferon-γ (IFN-γ, *P* = 0.002) (Fig. 1F), indicating that the altered NK1/NK2 balance results in a more pro-inflammatory NK repertoire. Collectively, these data show an imbalance in NK1/NK2 cells in peripheral blood of patients with BCR-UV and a skew towards a more proinflammatory phenotype.

### PBMC scRNA-seq identifies altered NK repertoire in Birdshot Chorioretinopathy

To allow characterization of the changes in peripheral blood NK cells in BCR-UV in an unbiased manner, we used single-cell RNA-sequencing (scRNAseq) of peripheral blood mononuclear cells (∼300K cells) of 24 BCR-UV patients and healthy controls (Fig. 2A and Supplementary Fig. 2A). Unsupervised clustering followed by uniform manifold approximation and projection (UMAP) and automated cell type annotation, identified an NK cell population (9,619 cells of cluster *C4*, Fig. 2B and Supplementary Fig. 2B) with an altered NK cluster structure in two-dimensional UMAP space in BCR-UV patients compared to healthy controls (Supplementary Fig. 2C). NK-specific *GZMB* (granzyme B), *KLRD1, GNLY, PRF1* (perforin), *NKG7* and *SH2D1B* (CD244 signaling) were among the most differentially upregulated genes in BCR-UV (Supplementary Fig. 2D). We extracted the NK cells from the PBMC scRNAseq data for further analysis.

**Figure 2.**
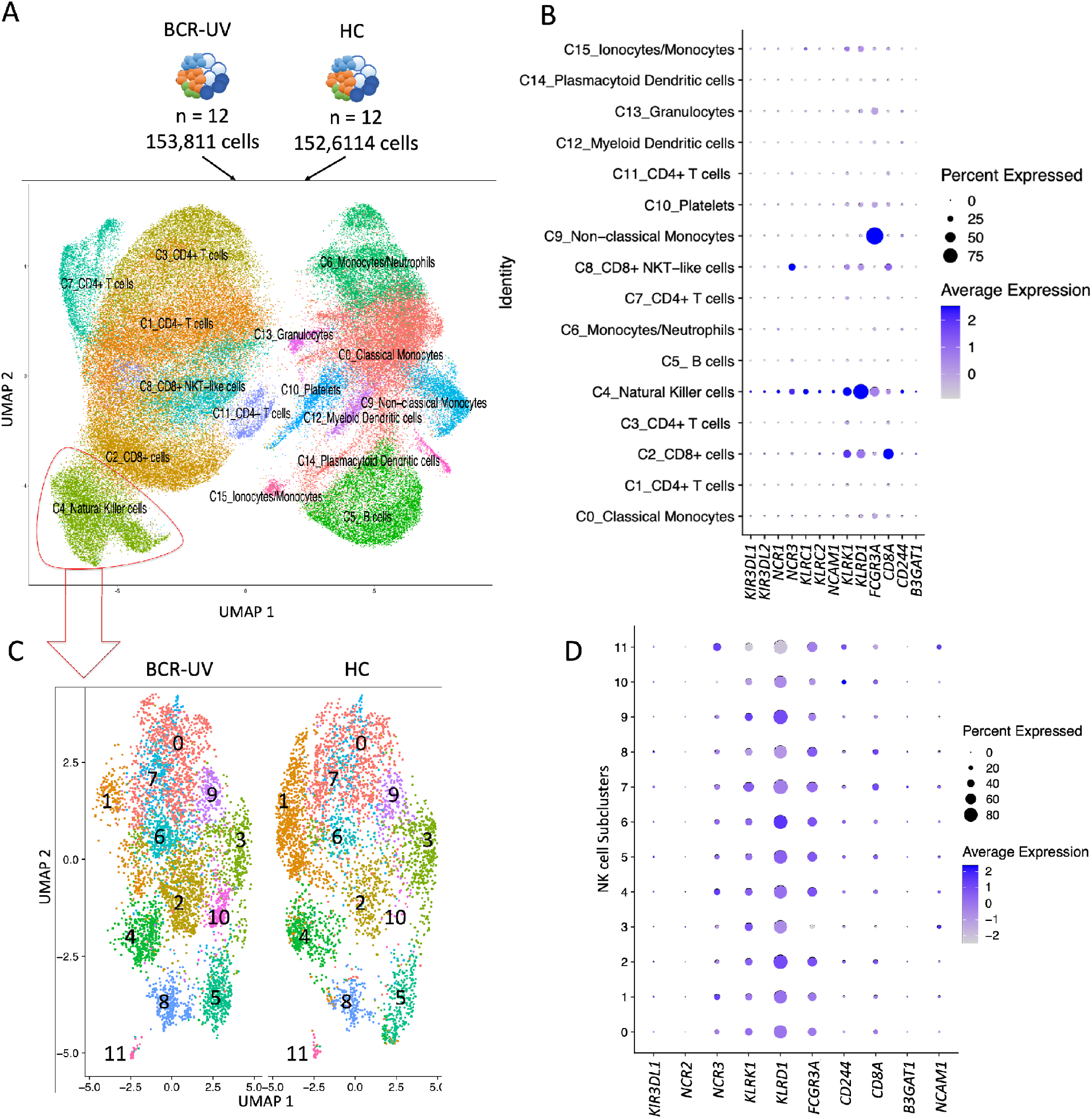
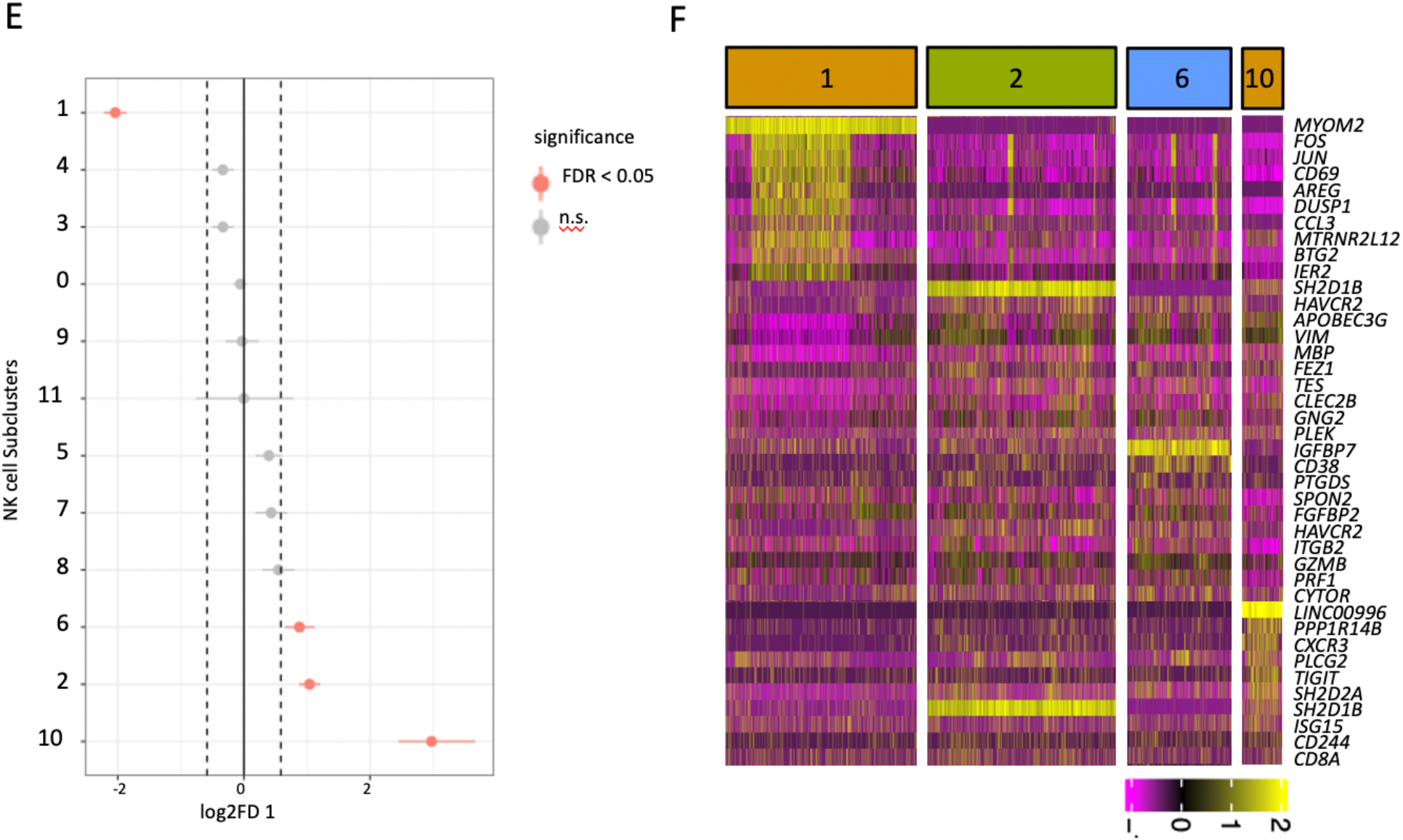
Single cell RNAseq analysis identifies transcriptional NK cell subsets associated with Birdshot Uveitis. **A**. UMAP plot of 306,425 cells from 12 birdshot uveitis patients (BCR-UV) and 12 healthy controls (HC). The identified lineage clusters are highlighted (color-coded). **B**. Dot plot showing the expression profile of NK surface marker encoding genes across the annotated cell clusters identified in *A*. **C**. UMAP plot of NK cells in BCR-UV and HC.The 12 transcriptional NK cell clusters are color-coded. **D**. Dot plot showing the expression profile of NK surface marker encoding genes in each of the clusters as identified in *C*. **E**. Proportion test analysis of NK cell clusters comparing between BCR-UV and Healthy controls. Significant differences at a false discovery rate of 5% [FDR < 0.05] and log2 fold difference = [Log2(FD)], are indicated in red. HC *n* = 12; BCR-UV *n* = 12. **F**. Heatmap of the gene expression profile of highly expressed genes (*n* = 10 unique genes) in clusters 1, 2, 6 and 10 of NK cells as identified in *E*.

Unsupervised clustering of the NK cell population revealed a high level of transcriptomic heterogeneity and the existence of 12 distinct clusters ranging from 57 cells (cluster 11) to 2,093 cells (cluster 0) in each cluster (Fig. 2C and Supplementary Fig. 3A). Gene expression levels of characteristic NK lineage surface markers revealed that these clusters expressed different levels of transcripts encoding NK activating receptors *CD244* and *CD8A*, as well as NK inhibitory receptors *KIR3DL1, KLRD1* and *B3GAT1* (Fig. 2D) which is compatible with an altered composition of functional NK subsets. At a false discovery rate of 5%, cluster 1 was significantly decreased (948 cells in HC vs 275 cells in BCR-UV) while clusters 2 (351 vs 862 cells in HC vs BCR-UV), 6 (208 vs 457 cells in HC vs BCR-UV) and 10 (25 vs 235 cells in HC vs BCR-UV) were significantly increased in frequency in BCR-UV patients compared to healthy controls (Fig. 2E). Clusters 1, 2, 6 and 10 were uniquely represented by the expression of *MYOM2, SH2D1B, IGFBP7* and *LINC00996* genes, respectively (Supplementary Fig. 3B). Cluster 1 also showed high expression of *DUSP1, FOS, JUN*, and *CD69* highly reminiscent of the gene expression profile of CD56^bright^ CD16^−^ NK cells [i.e., NK2]^20^, which corroborates our findings by major lineage flow cytometry (Fig. 2F). In contrast, the increased clusters 2, 6 and 10 expressed lower levels of *NCAM1* (CD56) (Fig. 2D) but high levels of *FCGR3A* (CD16) and the surface co-receptor encoding *CD8A* (CD8 Antigen, Alpha Polypeptide) (Fig. 2D), which suggests that clusters 2, 6, 10 are subpopulations of the bulk population of CD8+ NK1 cells.

We further observed that cluster 10 showed high *CD244* (Fig. 2D, F) and clusters 2 and 10 displayed enrichment of CD244 binding *Src homology 2* (SH2) domain-encoding genes *SH2D2A* and *SH2D1B*, which control signal transduction through the surface receptor CD244^29^ (Fig. 2F). This implicates active CD244 signaling in these NK clusters. Other highly expressed activation-associated genes in these sub-clusters include *TNFRSF18* (also known as *GITR*) and *ISG15* (Interferon-Stimulated Protein, 15 kDa) (Fig. 2F, Supplementary Fig. 3C, D). In summary, these results indicate expansion of activated CD8+ NK1 subpopulation characterized by high levels of CD244-signaling molecules in the blood of patients with BCR-UV.

### High-dimensional cytometry reveals accumulation of a CD8^bright^CD244^bright^ subset of CD16+ NK cells in the circulation of Birdshot Chorioretinopathy patients

We wished to validate our scRNAseq findings using a 12-marker panel (Supplementary Fig. 4A) flow cytometric phenotyping of the blood NK cell repertoire. Unbiased cell clustering considering the surface marker phenotypes by *FlowSOM* discerned 12 NK cell clusters (Fig. 3A). As expected, the majority of the clusters were CD56^dim^ CD16+ NK1 (cluster 0-2, 5-11) and a minor population was CD56^bright^ and CD16^−^ (NK2, *cluster* 3, 4) (Supplementary Fig. 4B). These 12 flow cytometry clusters broadly intersected with the NK clusters detected by the scRNAseq analysis: scRNAseq clusters 1 and 3 (Fig. 2C) are represented by the flow cytometry clusters 3 and 4 (Fig. 3A), all of which are CD56^bright^ population. The remaining 10 clusters in both scRNAseq and flow cytometry analyses are CD56^dim^ and CD16+ populations. Differential cluster abundance analysis revealed that clusters 4 and 5 were significantly reduced in the BCR-UV, while cluster 0 was significantly increased (Fig. 3B, C). The NK2 cluster 4 was further defined by high expression of CD336 and CD94 whereas the expanded NK1 cluster 0 was defined by high co-expression of CD8 and CD244, in line with our scRNA-seq data. We further found that cluster 0 expressed surface markers CD314 (NKG2D) and CD337 (NKp30), but not CD57 (Fig. 3D, E and Supplementary Fig. 4E). Principal component analysis supported that cluster 0 was the most distinguished cluster by high co-expression of CD8a and CD244 (Fig. 3E).

**Figure 3.**
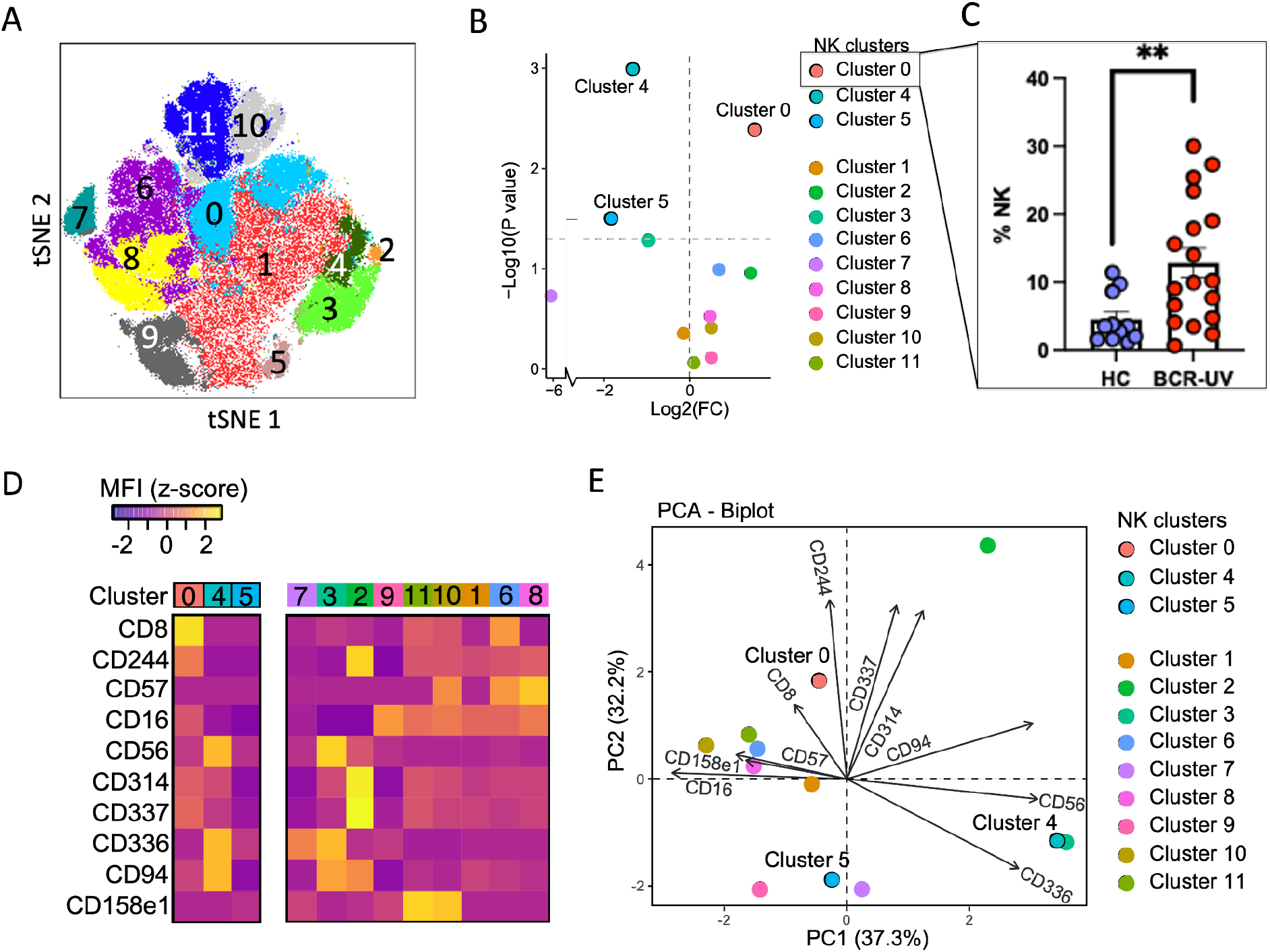
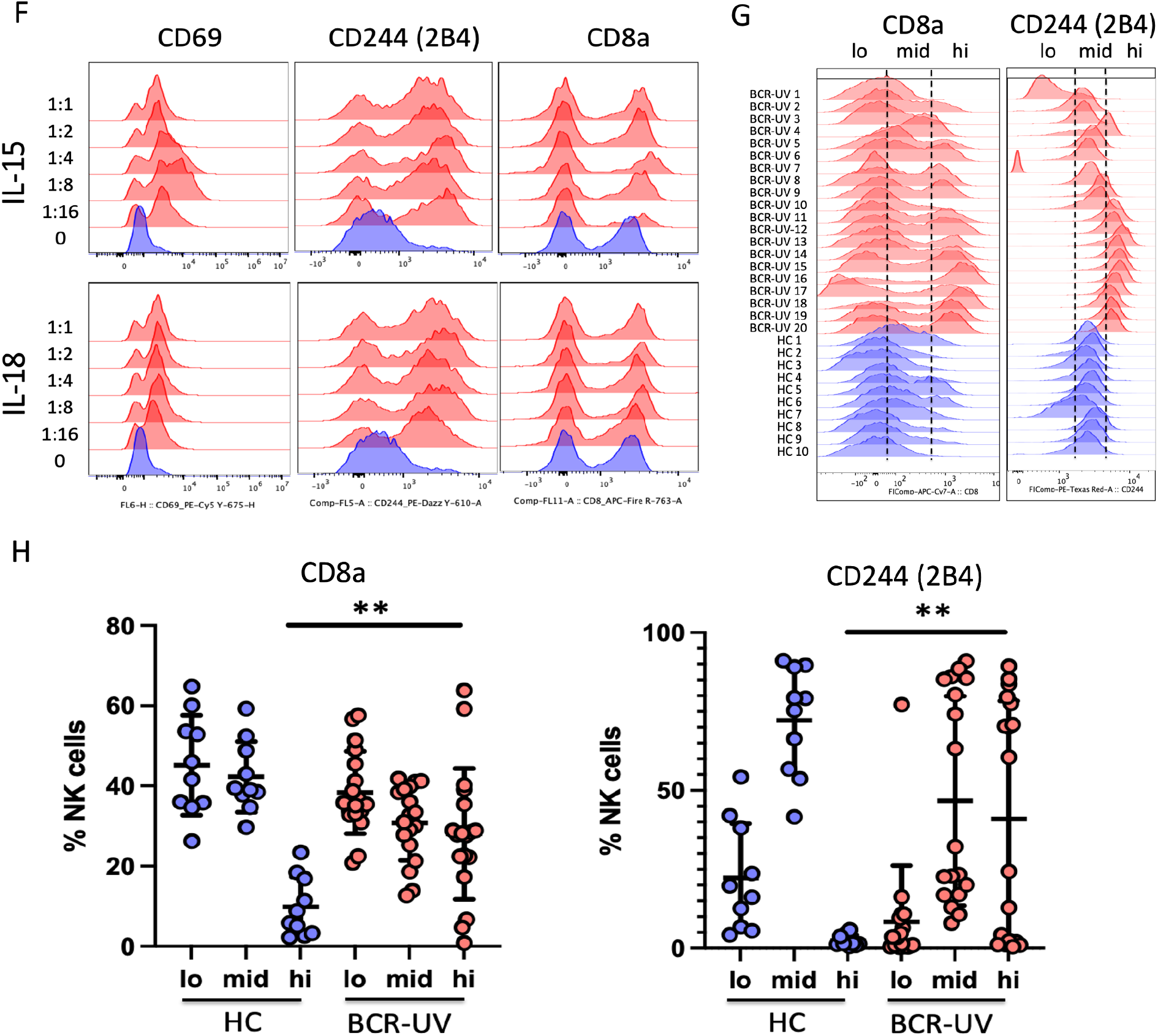
CD244^bright^ CD8^bright^ NK cell subset is significantly expanded in peripheral blood of Birdshot uveitis patients. **A**. t-SNE plot of flow cytometry data from Healthy control (HC) *n* = 10, Birdshot uveitis (BCR-UV) *n* = 18. Clusters identified by FlowSOM analysis (*n* = 12 clusters) are shown. **B**. Proportion test analysis of the NK clusters identified in *A*. Significantly different clusters 0, 4, and 5 are highlighted. **C**. Scatter plot of the frequency of cells of cluster 0 in peripheral blood in patients versus controls. **D**. Heatmap showing the mean fluorescent intensity (MFI) of surface markers in NK clusters. **E**. Principal component analysis (PCA) of NK clusters based on expression of surface markers. **F**. Histograms of the surface expression of CD69, CD244, and CD8 after 24 h stimulation of peripheral blood NK cells with recombinant human IL-15 and IL-18 proteins (dilution indicated). **G**. Histograms and corresponding **H**. frequency plots of CD8a and CD244 expression in circulating NK cells of Healthy controls (HC, blue) and Birdshot patients (BCR-UV, red). HC, *n* = 10; BCR-UV, *n* = 18.

We determined by *in vitro* culture that CD244 expression but not CD8 was upregulated in NK cells by restimulation with IL-15 and IL-18, indicating that CD8a+ CD244+ cells may represent activated CD8+ NK cells (Fig. 3F). To validate these findings, we manually gated the NK cell population based on the relative expression of CD8a and CD244 and divided the NK cells into three categories: CD8a^bright^/CD244^bright^ (hi), CD8a/CD244 ^medium^ (mid) and CD8a/CD244 ^low^ (lo) expressing cells. Quantitative analysis demonstrated that the CD8a^bright^/CD244^bright^ population was significantly more abundant in BCR-UV compared to that of healthy controls (*Student’s t-test, P* < 0.01) (Fig. 3G, H and Supplementary Fig. 4C, D). Together, these data show that CD8a^bright^/CD244^bright^ NK1 cells are enriched in the blood of BCR-UV patients.

### CD8a^bright^ CD244^bright^ cytotoxic NK cell frequency correlates with disease activity

Finally, we were interested to determine the dynamics of the newly identified NK cell subset in BCR-UV patients over the course of the disease. We had the opportunity to analyze samples taken prior to commencing therapy, during, and upon achieving clinical quiescence (according to the SUN criteria^30^) following treatment with systemic immunomodulatory therapy (1-year follow-up) (Fig. 4A, Supplementary Fig. 5A). We assessed the expression of CD8a and CD244 in the CD56^dim^CD16+ NK1 cell population in this cohort by flow cytometry. The mean fluorescent intensity (MFI) of surface expression for both CD8a and CD244 was elevated in patients with active BCR-UV and decreased over the course of treatment (Fig. 4B and Supplementary Fig. 5B). Similarly, the CD8a and CD244 double positive cells within CD56^dim^CD16+ NK population were significantly increased in patients with active BCR-UV and gradually decreased with one year of treatment and normalized to the frequency observed in healthy controls (*P < 0*.*05*) (Fig. 4C, D). In conclusion, these results show that CD8a^bright^/CD244^bright^ NK1 cells are expanded during active uveitis in BCR-UV patients but decrease upon successful systemic immunomodulatory treatment and clinical remission, compatible with the interpretation that CD8a^bright^/CD244^bright^ NK1 cells are a pro-inflammatory NK subset that are likely to be involved in the underlying disease mechanism.

**Figure 4.**
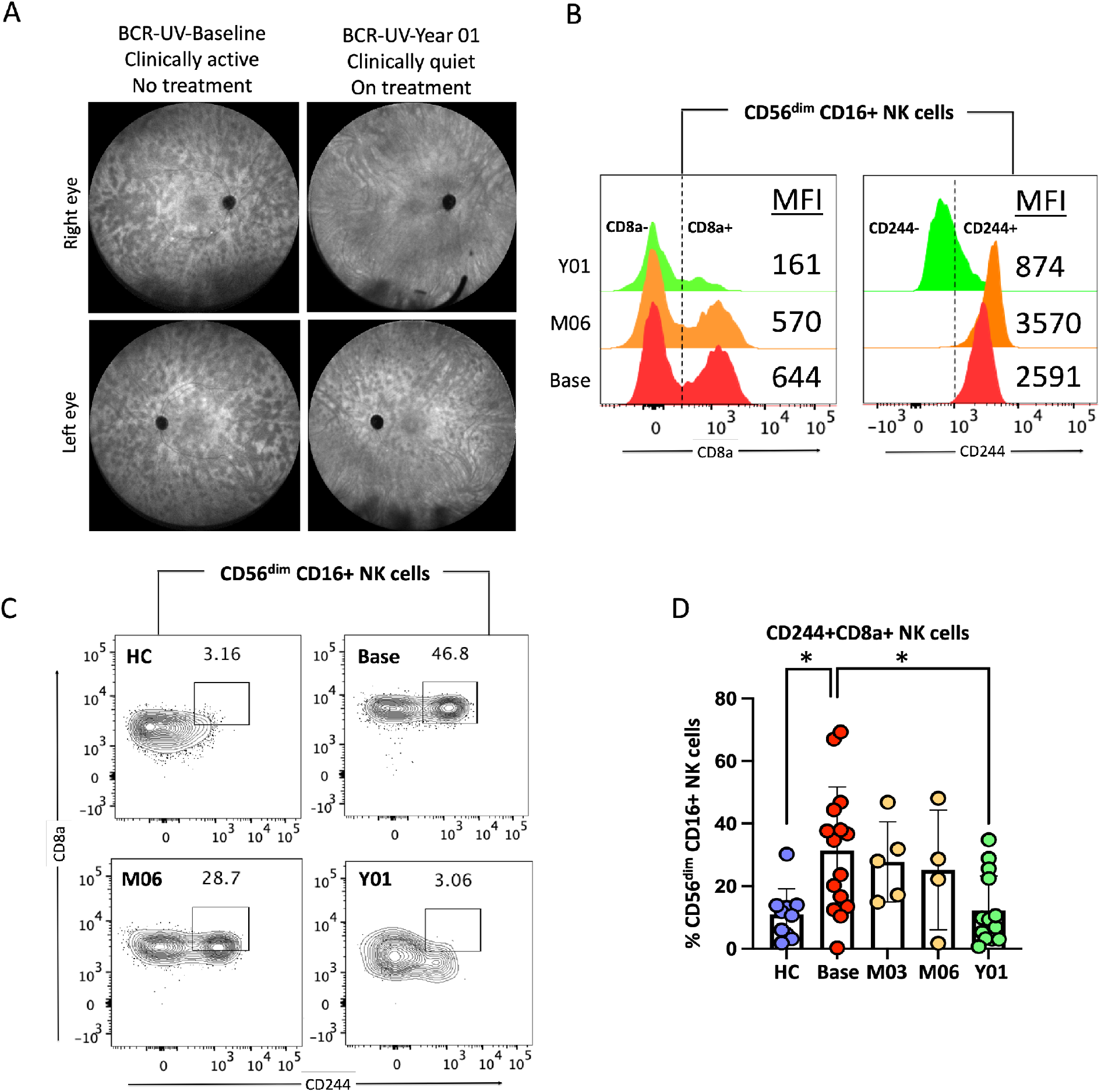
Response to systemic immunomodulatory therapy is accompanied by normalization of circulating CD244+CD8+ NK cells in patients. **A**. Representative Fluorescein angiography (FA) image the left and right eye of a Birdshot uveitis (BCR- UV) patient at baseline and after 1 year of systemic immunomodulatory therapy. **B**. Histograms are showing the expression of CD8a and CD244 proteins in circulating CD56dimCD16+ NK cell surface of healthy control (HC), BCR-UV patient at the baseline (Base) and on month 6 (M06) and year 1 (Y01). Values within the box indicate the mean fluorescent intensity (MFI) of CD8a and CD244 proteins. **C**. Representative flow plots show staining of CD8 and CD244 proteins within CD56dimCD16+ NK cells in the blood of healthy control (HC), BCR-UV patient at baseline with no treatment (Base), and longitudinally at follow-up visits at month 6 (M06) and year 1 (Y01) after treatments. **D**. Frequencies of CD244 and CD8 double positive cells within CD56dimCD16+ NK population in age-matched healthy control (HC) and in BCR-UV at baseline (Base) and treatment follow up visits. HC, *n* = 10; Baseline, *n* = 15; Month 03, *n* = 4; Month 06, *n* = 4 and Year 01, *n* = 15.

## DISCUSSION

In this study, we conducted deep molecular phenotyping of peripheral blood NK cells and identified altered changes in NK1/NK2 subsets in peripheral blood of BCR-UV patients. We found expansion of a CD8a^bright^ CD244^bright^ NK1 subset in BCR-UV patients that decreased upon successful treatment with systemic immunomodulatory therapy.

Our findings also corroborate previous reports on the decreased CD56^bright^ NK cells (NK2) in BCR-UV^17^ and the normalization of CD56^bright^ NK cell abundance upon immunosuppressive treatment of non-infectious uveitis^18^. In other MHC-I associated conditions, such as HLA-B51-associated Behcet’s disease (BD) and HLA-B27- associated ankylosing spondylitis (AS), NK1/NK2 changes have been reported. In BD, total NK cells are increased in frequency in blood^31, 32^ and produce increased IFN-gamma and TNF-alpha^33, 34^, which we also demonstrate in BCR-UV. In AS, the number of CD56^dim^ CD16+ subset of NK cells (NK1) in the peripheral blood is increased,^35, 36, 37^ which is in line with our finding of elevated CD16+ CD56^dim^ NK cells in BCR-UV. Here, we add to these previous observations that the decline in CD56^bright^ regulatory NK cells in BCR-UV is accompanied by a concomitant expansion of CD56^dim^ CD16+ NK cells, that also express the alpha-chain of the CD8 co-receptor for HLA class I^38, 39^, advancing the understanding of NK cell dynamics in ocular inflammatory disease. The subset of CD8+ NK cells (also known as “NK8^+^” cells^28^) make up approximately half of the blood NK cells^40^ and are found in both the CD56^dim^CD16+ and CD56^bright^CD16+ NK subsets (Supplementary Fig. 6A). Note that in contrast to CD8+ T cells, which express the alpha and beta chain of CD8, NK8^+^ cells only express the alpha chain of CD8 and mouse NK cells do not express CD8 at all^38, 39^.

NK8^+^ cells are considered to be functionally distinct and produce more IFN-gamma and TNF-alpha compared to CD8^−^ NK cells^41^. Accordingly, our data show that the NK8^+^- enriched population in patients secretes more IFN-gamma and TNF-alpha compared to healthy controls. In addition, the activation marker CD69 was increased in patients (Supplementary Fig. 6B) and was induced upon treatment with IL-15 or IL-18 (Fig. 3F).

This is significant because cross-linking of CD8 on NK cells induces increased expression of the activation marker CD69 at the cell surface^42^. Of interest, elevated frequency of peripheral blood NK8^+^ cells with homing marker CXCR3 is associated with an increased risk for Type 1 diabetes, a T-cell mediated autoimmune condition ^43^. The most expanded NK8^+^ subpopulation identified by scRNAseq (cluster 10) in BCR patients was also characterized by high *CXCR3* expression (Supplementary Fig. 3C). Whether this NK8^+^ subset (i.e., cluster 10) may directly contribute to eye inflammation in BCR-UV, remains to be determined. Alternatively, the NK8^+^ skewing may be a reflection of diminished regulatory capacity in the NK cell compartment. NK cells are required to suppress CD8+ T cell autoimmunity^44^ which is attributed to the negative immunoregulation of activated T cells by CD56^bright^ NK cells^45^. As shown in this study, CD56^bright^ NK cells are diminished in circulation of NIU patients, including BCR-UV patients. This makes it tempting to speculate that proinflammatory skewing of the NK cell population diminishes control of autoreactive CD8+ T cell immunity directed towards the eye in BCR-UV.

Detailed immunoprofiling by scRNAseq and flow cytometry revealed that expanded NK8^+^ cells in BCR-UV co-express high levels of other functional receptors, including CD244, which we demonstrate is a marker for activated NK cells. CD244 [or SLAMF protein 2B4] is an immunoregulatory surface receptor expressed by NK cells^46, 47^. Interaction of CD244 on NK cells with it’s ligand CD48 expressed by other cells causes the intracellular domain of CD244 to recruit proteins EAT2 and TSAd encoded by *SH2D1B* and *SH2D2A*, which were also associated with BCR-UV. The concomitant expression of *TNFRSF18* (Tumor Necrosis Factor Receptor Superfamily, Member 18) and *ISG15* (Interferon-Stimulated Protein, 15 kDa) levels in the novel NK subset supports the ‘inflammatory’ phenotype of this NK subset in BCR-UV patients. This is also supported by flow cytometry analysis that revealed expression of the activating receptor NKG2D (CD314) and the cytotoxicity receptor (NCR) NKp30 (CD337)^48^ by the CD8+ CD244+ NK cells in BCR-UV.

We showed that the CD244^bright^ NK8^+^ cells correlate with disease activity during longitudinal monitoring of patients treated with systemic immunomodulatory therapy. The NK8^+^ cell frequency has previously been shown to correlate with clinical parameters in several conditions, including HIV-1 and multiple sclerosis (MS)^28, 41^. More specifically, in MS, the NK8^+^ cell frequency was predictive of the relapse rate in a longitudinal cohort^28^. It would be interesting to use flow cytometry analysis of this cell subset to see if its abundance can be used to predict treatment outcome or clinical course in advance (measured at diagnosis).

In conclusion, using complementary immunophenotyping platforms, we identified an expanded CD8a^bright^ CD244^bright^ population of circulating NK cells in BCR-UV whose abundance reflects inflammatory disease activity. Better understanding of the molecular underpinnings of BCR-UV and its relation to clinical outcome may pave the way towards implementation of more effective personalized therapeutic approaches in ocular inflammatory diseases.

## METHODS

### Patients

This study was conducted in compliance with the Declaration of Helsinki and ethical principles regarding human experimentation. All samples were obtained under a *National Institutes of Health* (NIH) *Institutional Review Board* (IRB) approved protocol (Uveitis/Intraocular Inflammatory Disease Biobank (iBank); NCT02656381). Informed consent was obtained from all enrolled participants. We recruited 139 adult patients (Supplementary Table 1) with non-infectious uveitis (NIU), including 18 patients with Birdshot Chorioretinopathy uveitis (BCR-UV) at the National Eye institute (NEI) outpatient clinic. In total, 80 healthy donors- majority (iBank IDs) of whom were screened for no personal or family history of autoimmune diseases, were recruited and served as unaffected healthy controls. NIU was classified and graded in accordance with the SUN classification^30^. All patients with BCR-UV were HLA-A29-positive confirmed by HLA typing. A retrospective review of patient charts, fluorescein angiography (FA), indocyanine green angiography (ICGA), and electroretinography (ERG) was performed in order to determine disease activity, clinical course and response to systemic immunomodulatory treatment. Patient demographics are summarized in Supplementary Table 1.

### Blood sample processing

Blood samples from patients and healthy controls are collected through venipuncture and all the samples were processed within 4 h of blood collection. Fresh whole-blood samples were directly used for flow cytometry analysis. PBMCs were purified by standardized density gradient isolation (Ficoll-Paque) and stored in liquid nitrogen until further use.

### Flow cytometry

Three mL of whole blood was incubated with 30 mL 1x RBC lysis buffer (BioLegend *#420391*) at room temperature for 15 minutes, centrifuged at 400 x g for 5 minutes and resuspended in 3 ml ACK lysis buffer (Lonza #*BP10-548E*) for 4 minutes at room temperature. Cell suspension was washed in a 30 ml FACS buffer (FB: 1x PBS w/o calcium and magnesium chloride + 2 % FBS + 2 mM EDTA + 0.01 % NaN_3_). In total, 1- 3×10^6^ cells were stained for 45 minutes following resuspension in 100 uL FACS buffer with Fc block (Human TruStain FcX™, Biolegend # 422302) with the following antibodies (Supplementary Table 2): Alexa Fluor® 488 anti-human CD3 Antibody (clone OKT3), Alexa Fluor® 700 CD16 (clone 3G8), APC anti-human CD57 Antibody (clone HNK-1), APC-Fire 750 CD14 (clone 63D3), APC-Fire 750 CD8 (clone SK1), Biotin anti-human CD19 Antibody (clone HIB19), Biotin anti-human CD20 Antibody (clone 2H7), Biotin anti-human CD3 Antibody (clone SK7), Biotin anti-human CD56 Antibody (clone HCD56), BV 421™ anti-human CD158e1 Antibody (clone DX9), BV 421™ anti-human CD19 Antibody (clone HIB19), BV 510™ anti-human CD20 Antibody (clone 2H7), BV 510™ anti-human CD56 Antibody (clone HCD56), BV 650™ anti-human CD314 Antibody (clone 1D11), BV 650™ anti-human HLA-DR Antibody (clone L243), FITC anti-human CD4 antibody (clone SK3), FITC anti-human CD94 antibody (clone DX22), PE anti-human CD336 Antibody (clone P44-8), PE/Cy7 anti-human CD337 Antibody (clone AF29-4D12), PE/Dazzle™ 594 anti-human CD244 Antibody (clone C1.7), PerCP/Cy5.5 anti-human CD3 Antibody (clone SK7). BV 605 Streptavidin and Pe/Cy7 streptavidin were used for secondary staining. The Pe/Cy7 Lin cocktail in monocyte/DC panel includes anti-human CD3, CD19, CD20 and CD56 Antibodies. Live/dead staining was carried out using Fixable Viability Dye eFluor® 455UV concomitantly with surface staining. Cells were fixed with Fixation Buffer (Biolegend #420801) and resuspended in 300 uL FACS buffer for acquisition on BD LSR Fortessa 1-3 days after staining.

Data were analyzed using FlowJo v10. For ex vivo restimulation, NK cells from a healthy donor were cultured with recombinant human IL-15 and IL-18 using five serial dilutions as indicated of ED_50_ concentrations. Cell-surface expression of CD69, CD244 and CD8 was measured using a BD LSRFortessa and analyzed by FlowJo V.10.

### Intracellular cytokine staining

For intracellular cytokine staining, cryopreserved PBMCs were thawed into warm RPMI/10% FBS, washed once in cold PBS and divided cells in two equal proportions. Protein Transport Inhibitor Cocktail (eBioscience #00-4980-03, 1/500 vol/vol) was added to each proportion. Cells were incubated with cell stimulation cocktail (eBioscience # 00- 4970-03, 1/500 vol/vol) for 4 hours at 37 degrees or kept under similar conditions without the stimulation cocktail. Cells were washed with cold PBS and resuspended in 100 uL FBS with Fc block (Human TruStain FcX™, Biolegend # 422302) followed by surface markers staining for 45 min. Cells were then washed once in cold PBS and incubated in fix/perm buffer (eBioscience FOXP3/Transcription Factor Staining Buffer, invitrogen #00-5523-00) for 20 min and then incubation with the following intracellular antibodies for another 30 min: BV510 anti-human Granzyme B (clone GB11), APC/Cyanine7 anti-human TNF-α (clone MAb11), PE/Dazzle 594 anti-human IFN-gamma (clone 4S.B3) and PE anti-human Perforin (clone B-D48).

### FlowSOM analysis

Live NK cells (Aqua L/D-CD3-CD20-CD56+) were gated from 18 BCR-UV patients sampled at baseline (i.e., at disease onset or relapse) and 10 healthy controls. NK cell fraction (range 5,011 to 42,358 NK cells) was down-sampled to 5,000 NK cells per donor using FlowJo DownsampleV3 plugin, concatenated and subjected to t-distributed stochastic neighbor embedding (*t-SNE*, iteration 1000, perplexity 30). The FlowSOM plugin in FlowJo was used to cluster cells into 12 meta-clusters (following 12 clusters identified in scRNAseq).

### PBMC single cell RNA sequencing

PBMCs from a total of 24 BCR-UV patients and healthy controls (Supplementary Table 3) were thawed quickly at 37°C and resuspended in RPMI media supplemented with 10% FBS. Approximately, 1,000-1,200 viable cells per microliter were loaded for capture onto the Chromium System using the v2 single-cell reagent kit (10X Genomics). Following capture and lysis, scRNA-Seq libraries were prepared from ∼16k cells using a 10X Genomics Chromium device and Chromium Single Cell 3’ Reagent Kit v3.1 according to manufacturer’s protocol. GEX (transcriptome) libraries were sequenced on a NovaSeq 6000 DNA sequencer (Illumina, Inc.) using V1.5 chemistry, generating ∼160M GEX reads per sample and ∼40M ADT reads per sample. The raw data was processed using RTA 3.4.4. Sequencing information is shown in Supplementary Table 4.

### Preprocessing of scRNAseq data

The raw fastq data were processed using *cellranger* v6.0.0^49^ with genome assembly GRCh38 (hg38), 3’ and 5’ assay chemistry SC3Pv3, and an expected cell count of 10,000 per sample (Supplementary Table 5). Further analysis of the single cell data was done using the *Seurat* v4.0.5^50^ package in the R v4.1.0 environment^51^. Cells with fewer than 300 genes and number of transcript counts less than 1,000 and more than 12,000, more than 8% mitochondrial and 40% ribosomal fraction were excluded from the dataset. Doublets were detected and removed using the *doubletFinder v3*.*0* package^52^ in R, set with an expected doublet level of 7%. Data were normalized using the “LogNormalize” method with the scaling factor set at 10,000. The variables, sample batch, percent mitochondria, percent ribosomes, transcript counts and gene counts, were regressed out using the ScaleData function.

### Dimension reduction, clustering and visualization

Cells were clustered using Principal Component Analysis (PCA) using the RunPCA function. The first 20 PCs identified (by Elbow method) were used in the ‘FindNeighbors’ (based on k-nearest neighbor (KNN) graphs) and ‘FindCluster’ (Louvain algorithm) functions in Seurat. RunTSNE and RunUMAP functions were used with “pca” as the reduction method, to visualize the data. The *FindAllMarkers* function was used to identify cluster specific markers. Cell type annotation was generated manually combining automatic annotation results from *ScType*^53^, with “Immune System” set as the tissue type, and *SCSA*^54^, with the whole database as reference. Outcomes from ScType and SCSA were used to curate cluster annotation and identify NK cells by plotting expression of lineage-specific genes. Differences in proportions of scRNAseq data between the groups were assessed using the “scProportionTest” package^55^, with number of permutations set at 10,000.

## Data Availability

All data produced in the present study are available upon reasonable request to the authors.

## Abbreviations

BCR-UV: birdshot chorioretinopathy uveitis;
NIU: non-infectious uveitis;
KIR: killer cell immunoglobulin-like receptors;
scRNAseq: single-cell RNA sequencing;
MFI: mean fluorescent intensity;
SUN: standardization of uveitis nomenclature.

## Data availability

All raw data and workflow are available as an open-source resource, with documentation.

## ACKNOWLEDGEMENT

We acknowledge the NIAMS Flow Core (James Simone, Kevin Tinsley, Jeffrey Lay), NEI Flow Core (Rafael Villasmil, Jyothi Priya Mandala) and Jenny McDowell at NISC for help with this project.

## Supplementary data for

**Figure S1.**
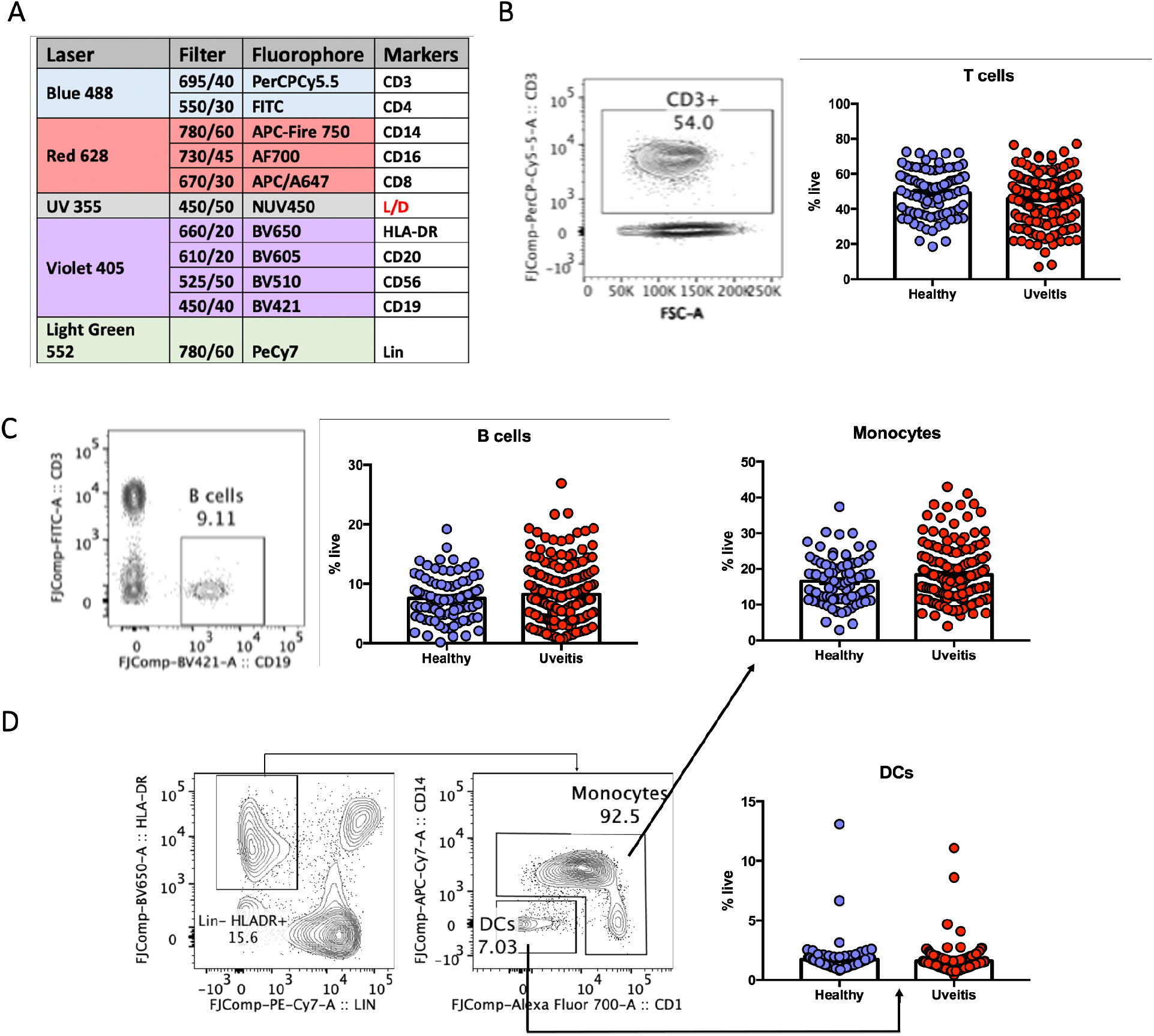
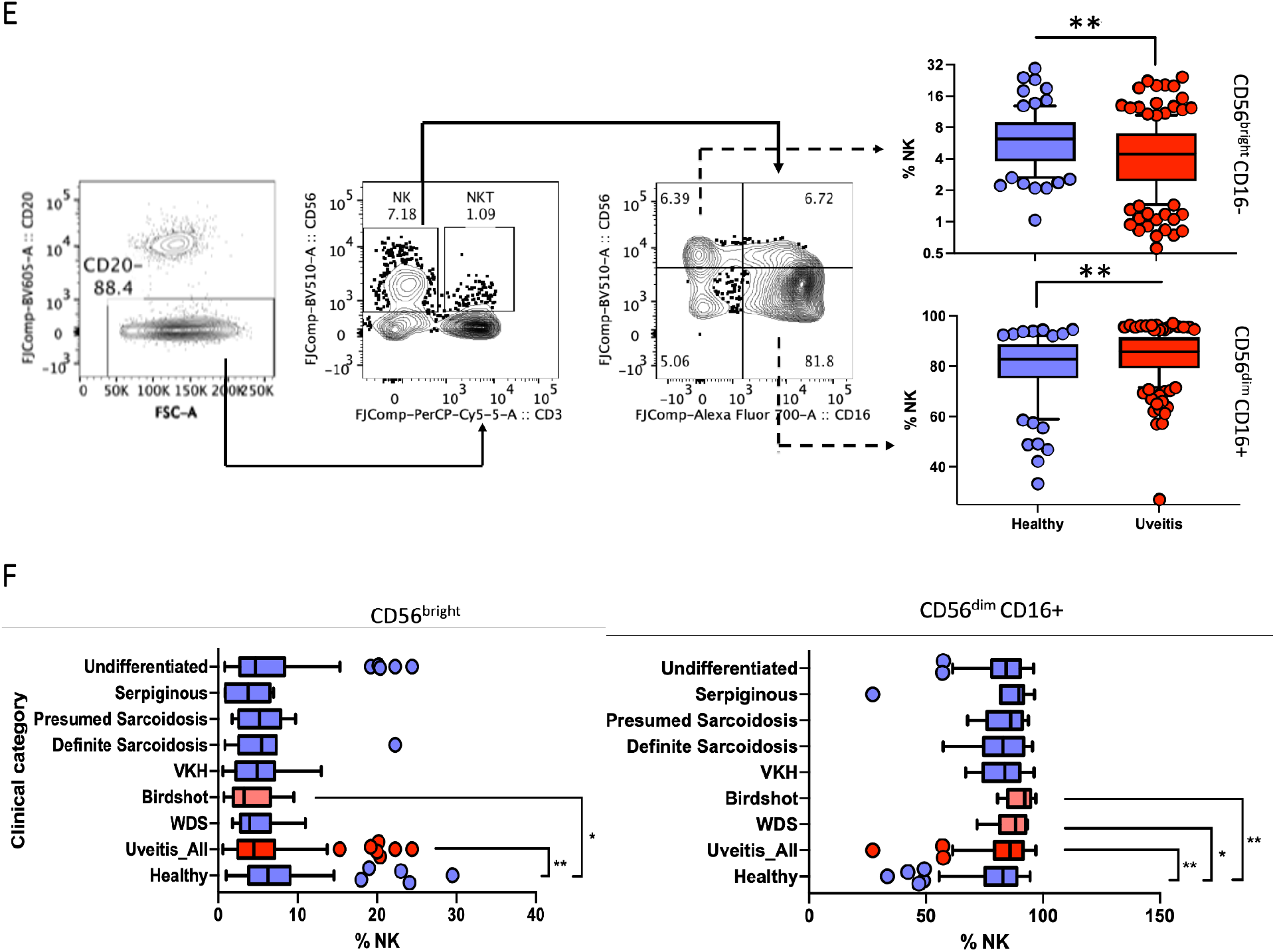
**A**. List of markers for interrogating major lineages of immune cells (10 markers, 6 major immune subsets) in peripheral blood of patients and healthy controls using flow cytometry. Representative plot and frequency of **(B)** T cells, **(C)** B cells, **(D)** monocyte and (DCs) and **(E)** NK cells and in the peripheral blood of uveitis patients (blue) and healthy controls are shown. There was no change in the frequency of T cells, B cells, DCs and monocytes, except the differences found in the NK cell subsets between the two groups. CD56highCD16- NK cells were significantly decreased while concomitantly CD56loCD16+ NK cells were significantly increased in uveitis cohort compared to the healthy controls. **F**. Flow-cytometry analysis of NK1 (CD56dim CD16+) and NK2 (CD56bright) NK cells (CD3^−^CD19^−^CD56^+^) in the fresh blood of different uveitis subgroups.

**Figure S2.**
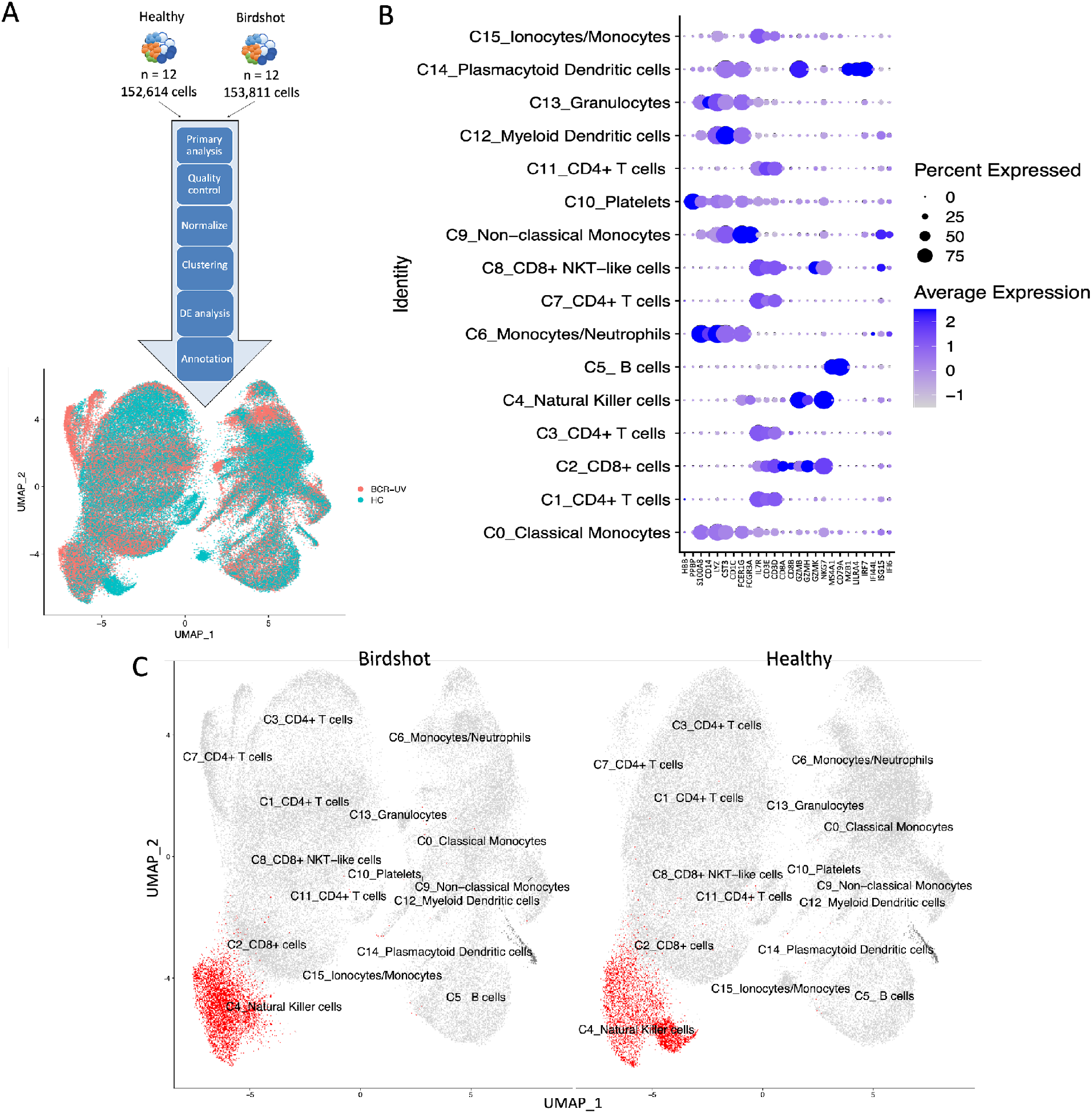
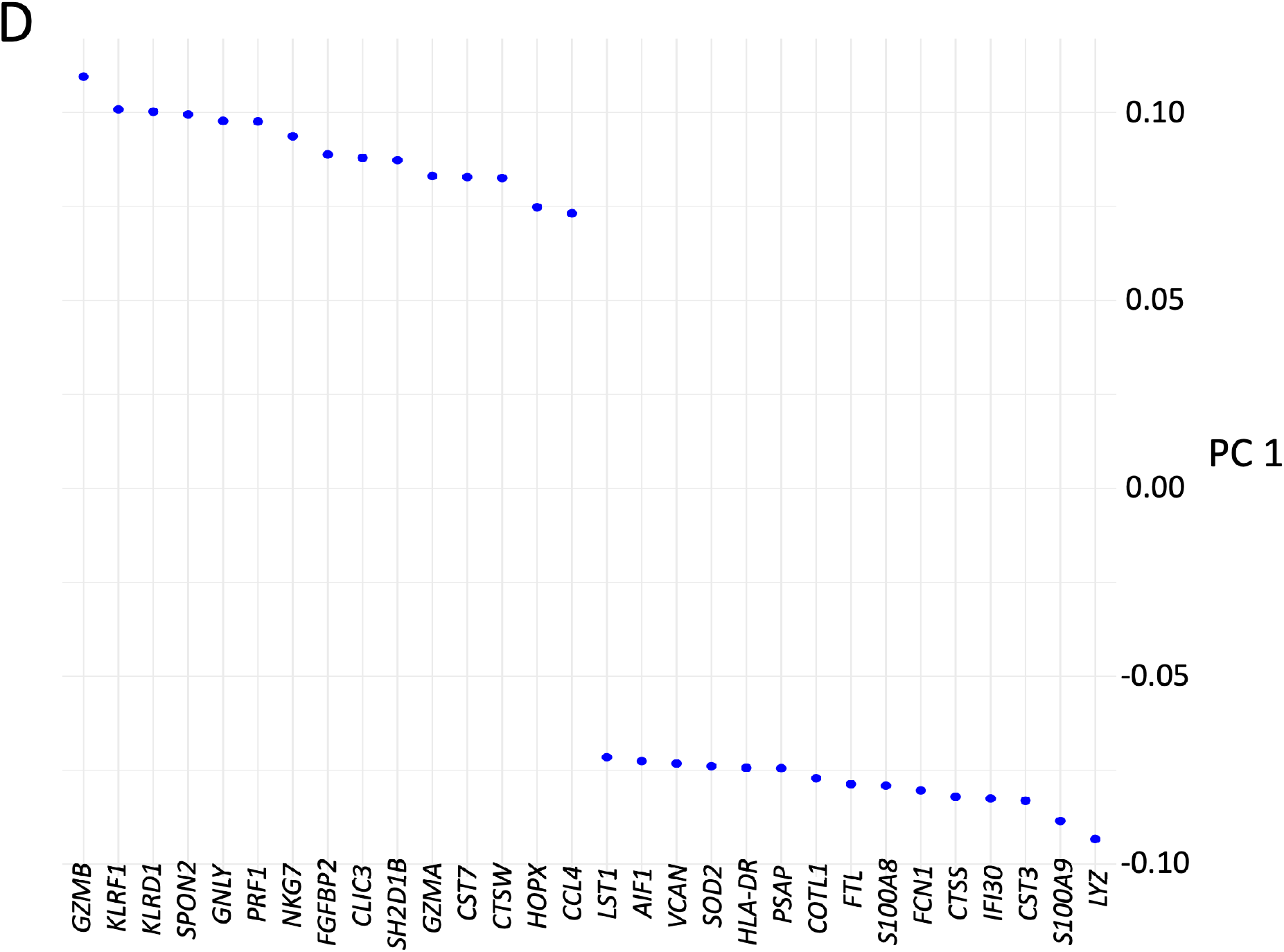
**A**. An overview of scRNAse analysis. A total of 306,425 cells from 12 Birdshot uveitis patients (Birdshot/BCR-UV) and 12 healthy controls (Healthy) were subjected to 10x Chromium single cell separation and sequencing and merged to a combined dataset. The bioinformatic analysis pipeline includes the Primary analysis, Quality control, Normalization, Clustering, Differential Expression (DE) analysis and Annotation of the combined dataset. The UMAP plot (top) shows the merged distribution of cells from Healthy (Green) and BCR-UV (Red). The UMAP plot (bottom) shows annotation of 16 clusters of cell lineages in the combined dataset. The putative identity of each cluster was manually annotated based on ‘scsa’ and ‘scType’ annotations and interrogating the top expressed genes in each cluster. **B**. Dot plot showing the expression profile of lineage-specific genes in the annotated cell clusters. **C**. The UMAP plot of single cell distribution is representing 16 clusters of cell lineages in the combined dataset of a total of 306,425 analyzed cells. The putative identity of each cluster was manually annotated. The NK cell clusters are identified in red color and the remaining cell clusters are presented in grey color. **D**. The top 30 most differentially expressed NK genes defined by principal component 1 in BCR-UV compared to HC.

**Figure S3.**
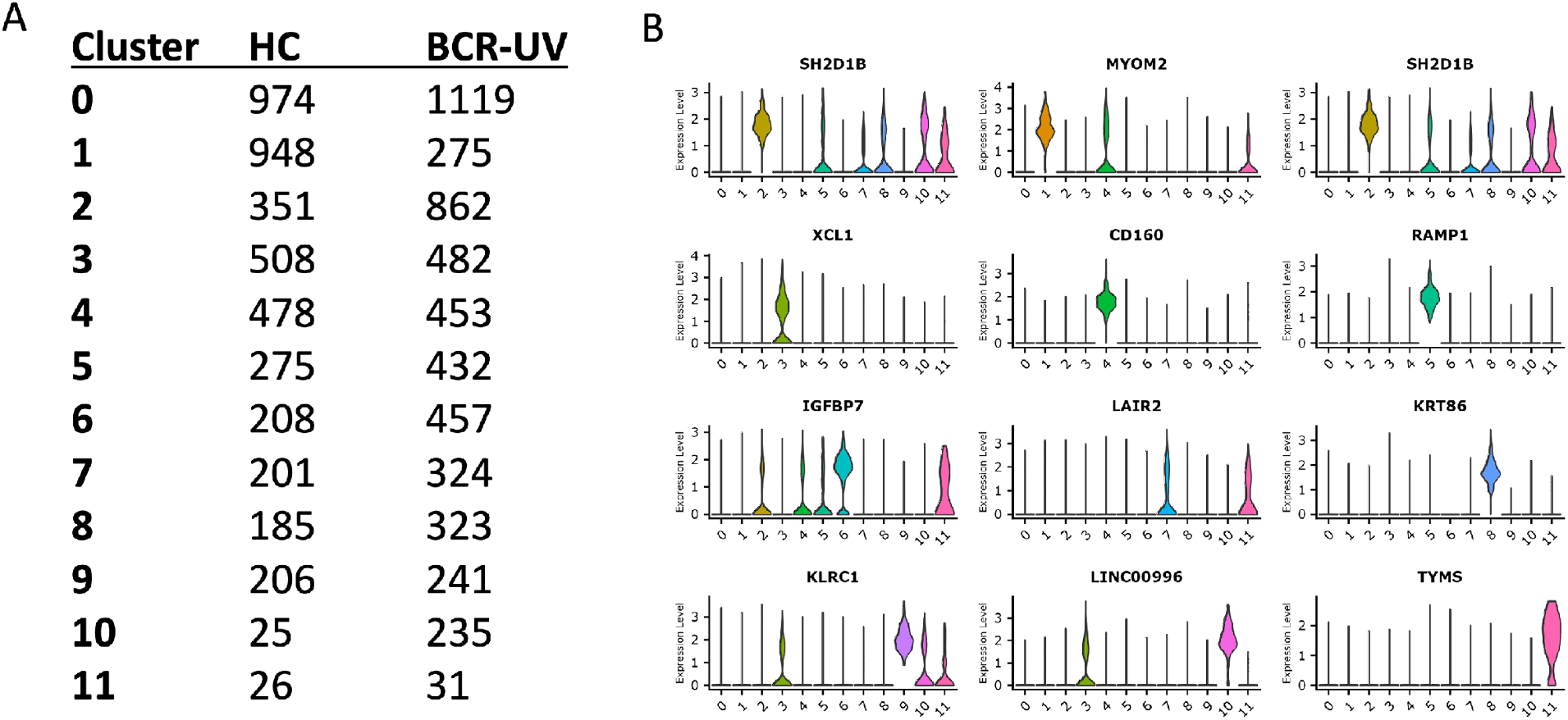
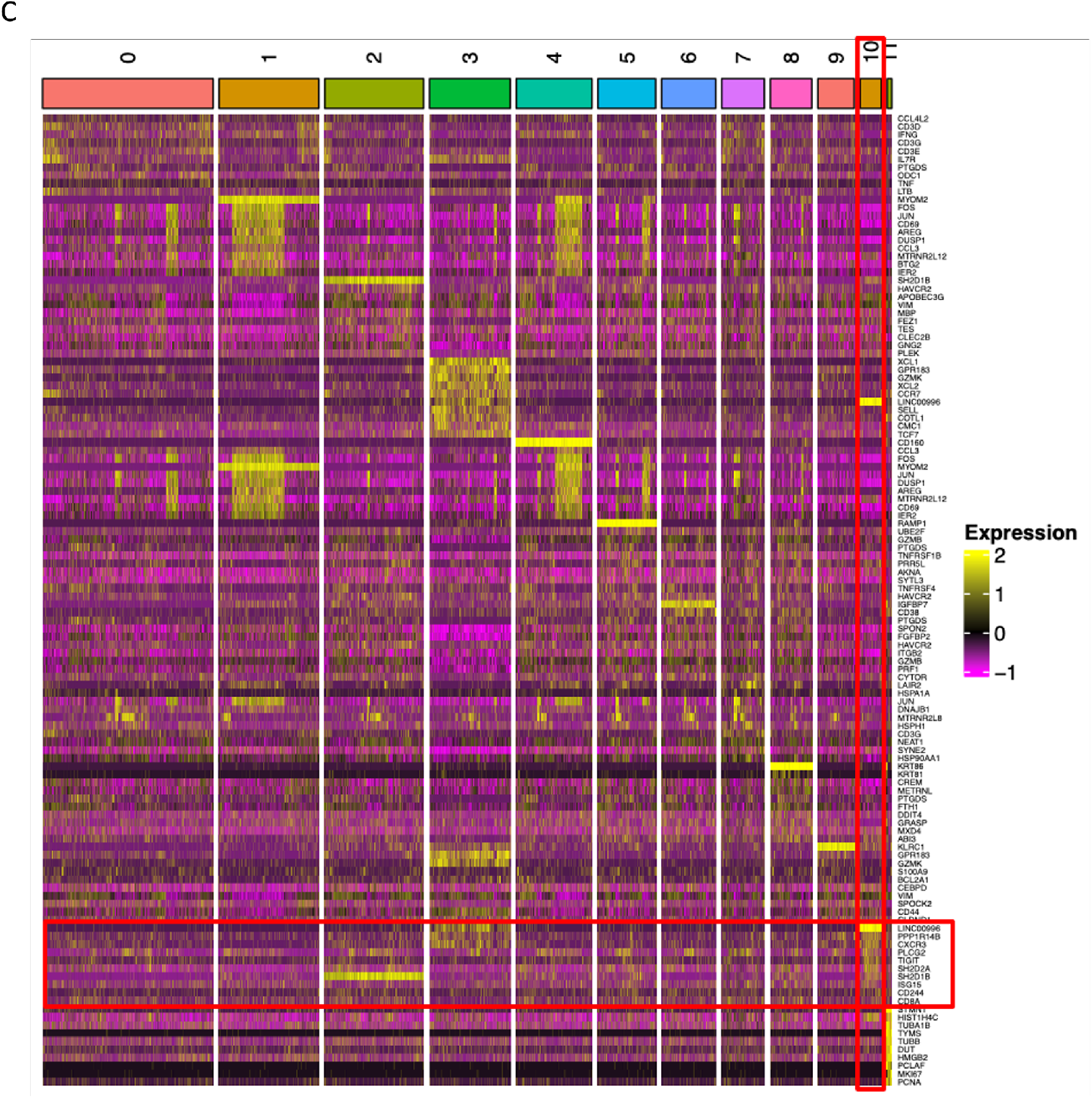
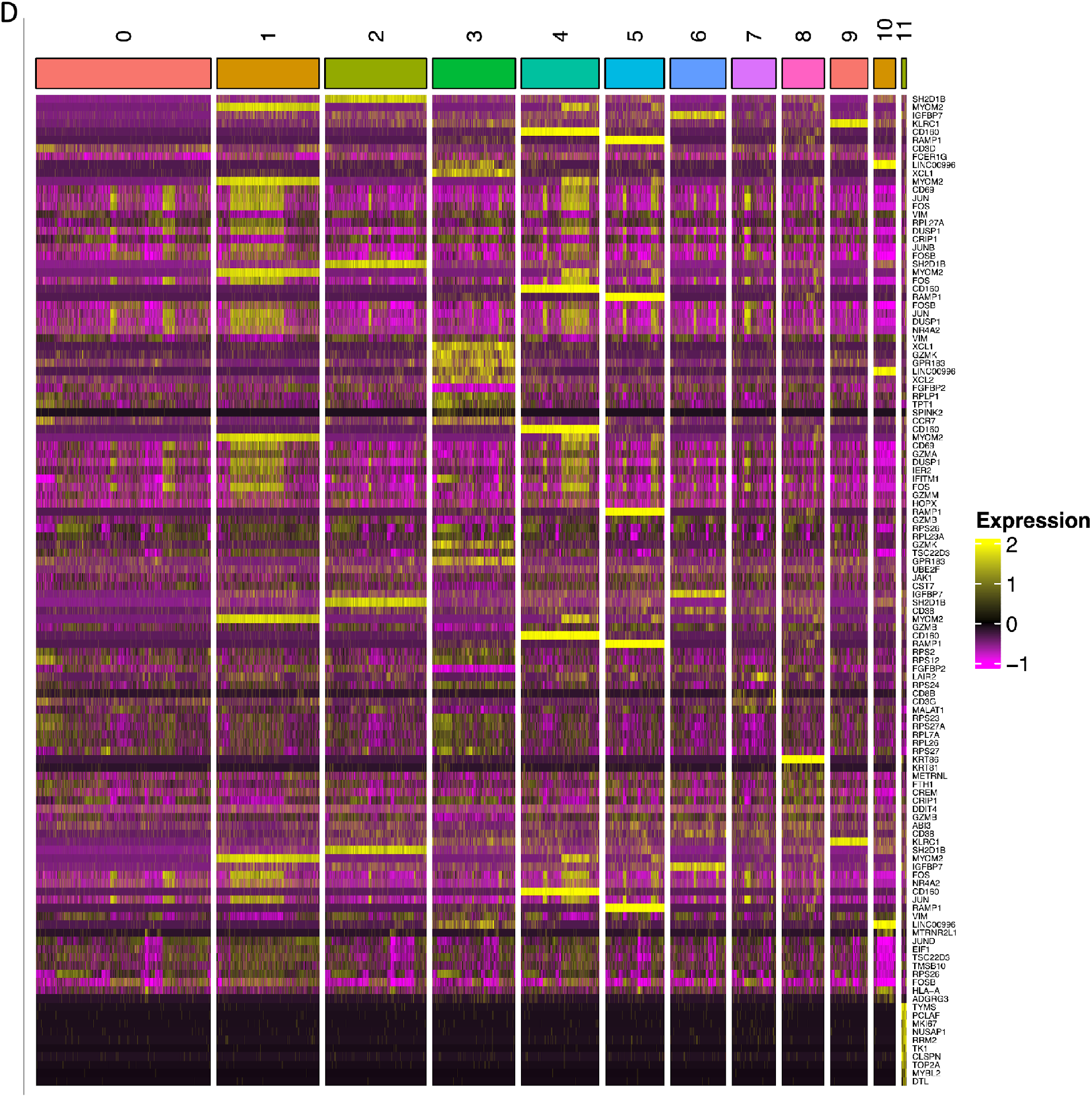
**A**. Tabular representation of total 12 NK-subclusters and associated cell count in 12 healthy controls (HC) and in 12 birdshot uveitis patients (BCR-UV). **B**. Violin plots representing the list of genes that are uniquely expressed in each of the 12 clusters of NK cell-subclusters. **C**. Heatmap representing the expression of top 10 highly expressed markers of each NK cluster. **D**. Heatmap representing the expression of top 10 differentially expressed markers of each NK cluster.

**Figure S4.**
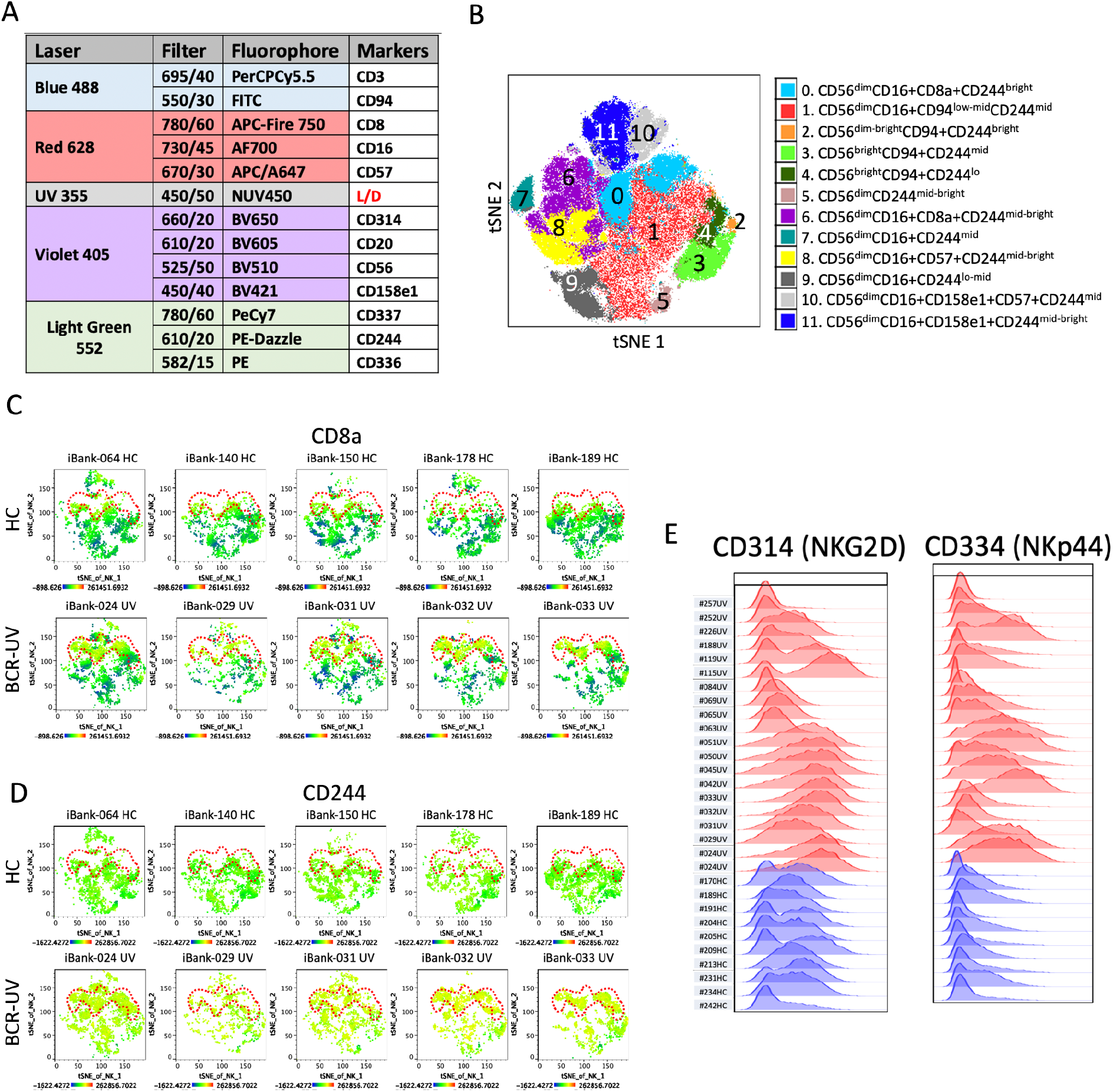
**A**. List of markers used to interrogate NK lineage cells in peripheral blood of BCR-UV patients and healthy controls using flow cytometry. **B**. t-SNE plot represents the FlowJo-based FlowSOM analysis to identify clusters based on NK cell surface markers from a combined dataset of healthy controls and Birdshot patients. Cluster 0 (CD56dim CD16+ CD8+ and CD244bright) is one out of 12 NK clusters that was elevated in Birdshot. Phenotypes of all 12 NK clusters are color coded and described. Healthy *n* = 11, Birdshot *n* = 18. **C**. Distribution of CD8a and **D**. CD244 surface protein expression is indicated in t-SNE plots of NK cells in five healthy controls (HC) and five birdshot uveitis (BCR-UV). Red dotted line captures the region where NK cells are present in BCR-UV but absent from HC. **E**. Histograms showing expression of CD314 (NKG2D) and CD344 (NKp44) in 10 healthy controls (blue) and 20 birdshot uveitis (red).

**Figure S5.**
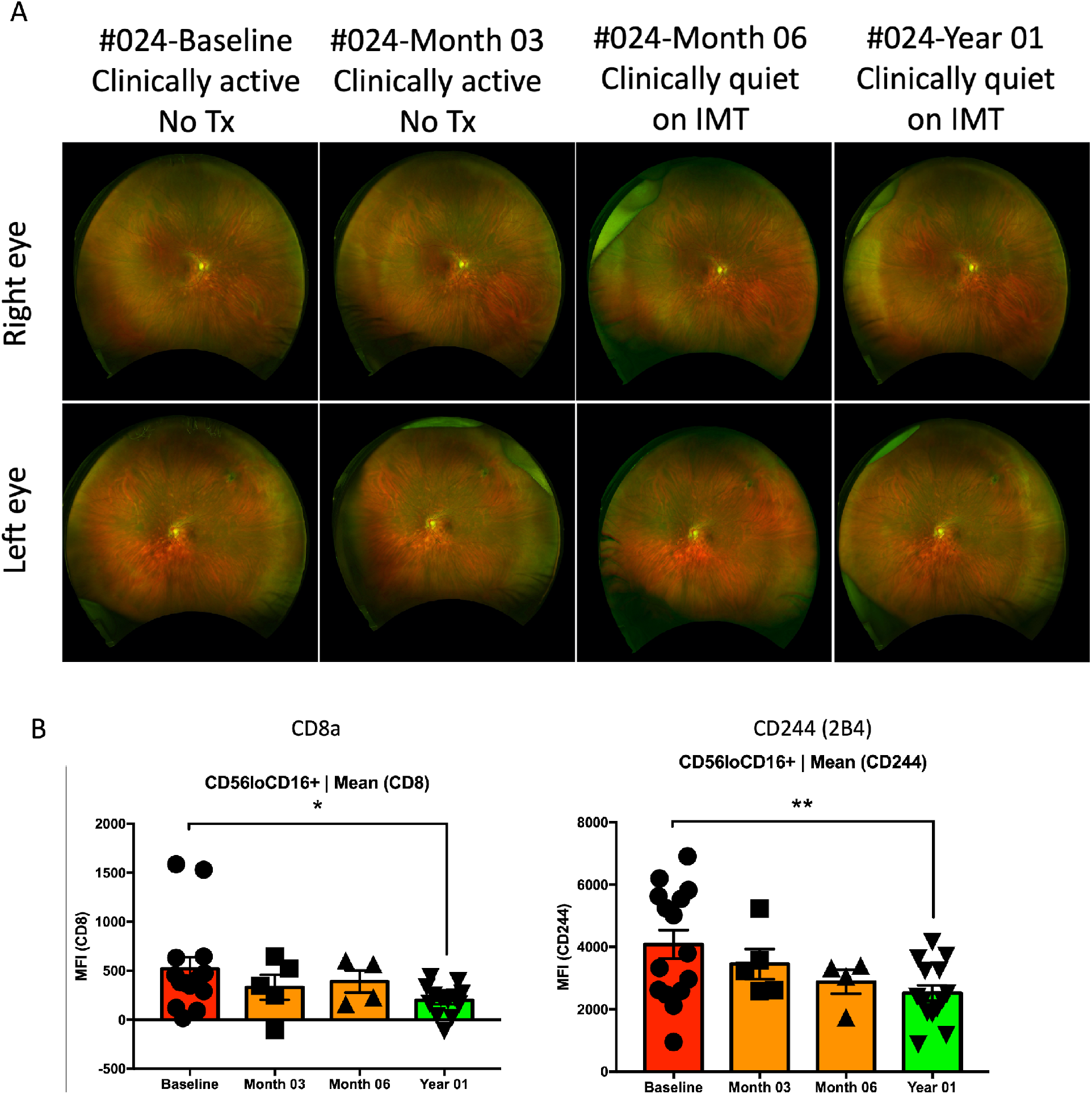
**A**. Fundoscopy examination of both eyes of a Birdshot uveitis patient (#024) at the Baseline, after Month 03, Month 06 and Year 01 of systemic immunomodulatory treatment (IMT). **B**. Bar plots indicate statistical significance on changes of CD8a and CD244 MFI within CD56loCD16+ NK cells from 15 different Birdshot uveitis patients from baseline to year 1 after treatment.

**Figure S6.**
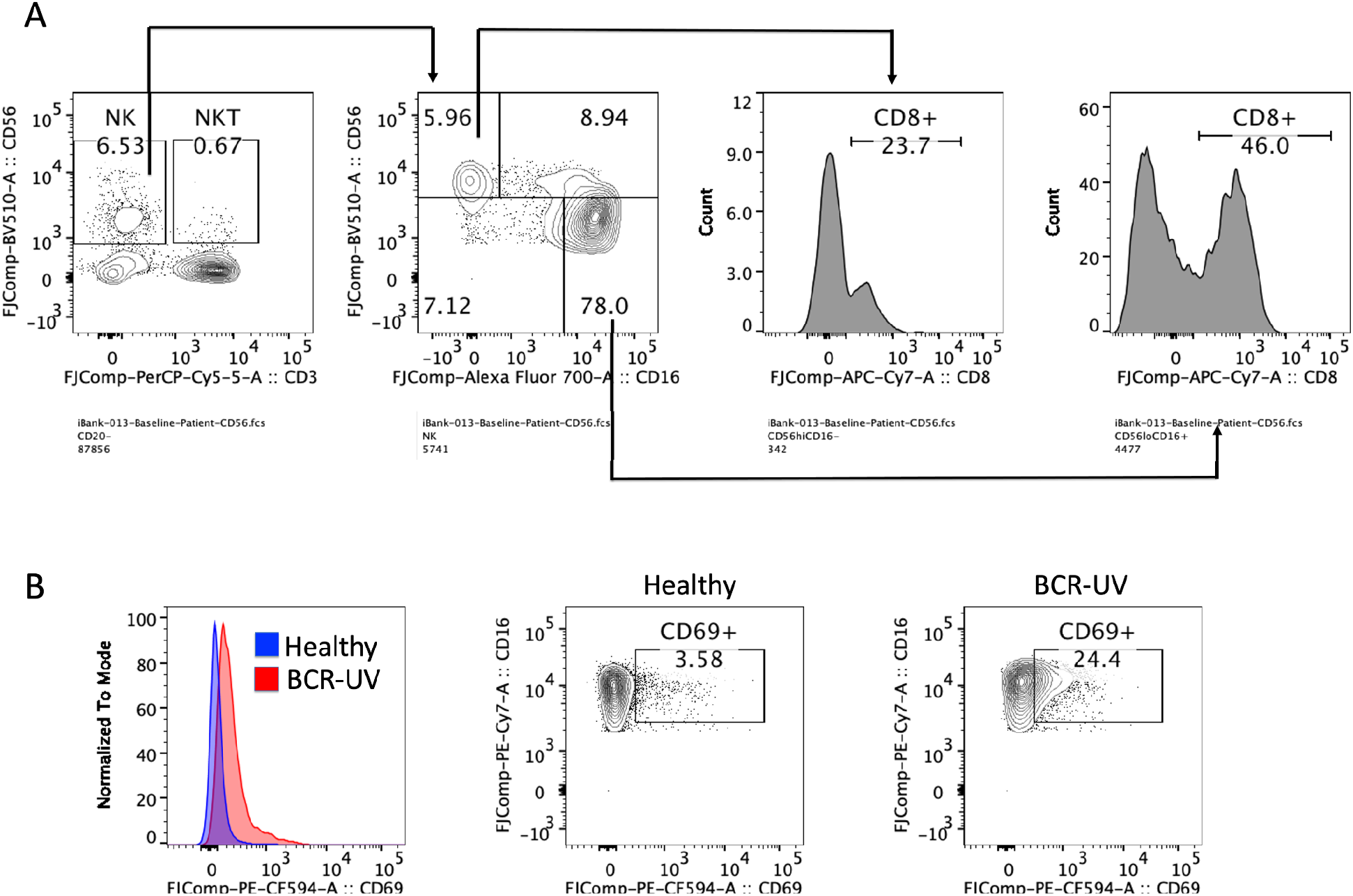
**A**. CD8 expression profiles in CD56bright and CD56dimCD16+ subsets of NK cells. **B**. CD69 expression in CD56dim CD16+ NK cells of healthy vs BCR-UV patients.

